# Effective Connectivity and Dopaminergic Function of Fronto-Striato-Thalamic Circuitry in First-Episode Psychosis, Established Schizophrenia, and Healthy Controls

**DOI:** 10.1101/2021.03.11.21253426

**Authors:** Kristina Sabaroedin, Adeel Razi, Sidhant Chopra, Nancy Tran, Andrii Pozaruk, Zhaolin Chen, Amy Finlay, Barnaby Nelson, Kelly Allott, Mario Alvarez-Jimenez, Jessica Graham, Lara Baldwin, Steven Tahtalian, Hok P Yuen, Susy Harrigan, Vanessa Cropley, Sujit Sharma, Bharat Saluja, Robert Williams, Christos Pantelis, Stephen J. Wood, Brian O’Donoghue, Shona Francey, Patrick McGorry, Kevin Aquino, Alex Fornito

## Abstract

Dysfunction of fronto-striato-thalamic (FST) circuits is thought to contribute to dopaminergic dysfunction and symptom onset in psychosis, but it remains unclear whether this dysfunction is driven by aberrant bottom-up subcortical signaling or impaired top-down cortical regulation. Here, we used spectral dynamic causal modelling (DCM) of resting-state functional magnetic resonance imaging (fMRI) to characterize the effective connectivity of dorsal and ventral FST circuits in a sample of 46 antipsychotic-naïve first-episode psychosis (FEP) patients and 23 controls and an independent sample of 36 patients with established schizophrenia (SCZ) patients and 100 controls. We found that midbrain and thalamic connectivity were implicated across both patient groups. Dysconnectivity in FEP patients was mainly restricted to the subcortex, with positive symptom severity being associated with midbrain connectivity. Dysconnectivity between the cortex and subcortical systems was only apparent in SCZ patients. In another independent sample of 33 healthy individuals who underwent concurrent fMRI and [^18^F]DOPA positron emission tomography, we found that striatal dopamine synthesis capacity was associated with the effective connectivity of nigrostriatal and striatothalamic pathways, implicating similar circuits as those associated with psychotic symptom severity in patients. Our findings thus indicate that subcortical dysconnectivity is salient in the early stages of psychosis, that cortical dysfunction may emerge later in the illness, and that nigrostriatal and striatothalamic signaling are closely related to striatal dopamine synthesis capacity, which is a robust risk marker for psychosis.

## Introduction

Dysfunction of fronto-striato-thalamic (FST) circuits linking the caudate and putamen with the prefrontal cortex is thought to be central to the emergence of psychotic symptoms [1–6]. Two such circuits are particularly relevant: 1) a ventral ‘limbic’ system involved in emotional and reward processing, which connects the orbital and ventromedial prefrontal cortices and subcortical limbic structures (e.g., hippocampus and amygdala) with the nucleus accumbens (NAcc); and 2) a dorsal ‘associative’ system, which links the dorsolateral prefrontal cortex (dlPFC) with the dorsal striatum, and subserves associative learning and executive functions [1, 7]. Feedback loops passing through the pallidum and the thalamus connect both circuits back to the cortex [1].

The striatum is a major target for dopamine projections from the ventral tegmental area (VTA) and substantia nigra (SN), with the ventral and the dorsal regions respectively forming part of the mesolimbic and nigrostriatal pathways [8]. Dysregulated dopamine signaling is proposed to contribute to psychosis onset by influencing the capacity of the striatum to filter and relay information to the thalamus, thus affecting broader FST function [9]. In-vivo positron emission tomography (PET) has revealed elevated presynaptic dopamine synthesis capacity, measured using 3,4-dihydroxy-6-[^18^F]-fluoro-L-phenylalanine ([^18^F] DOPA), in the dorsal striatum of at-risk groups, especially in individuals who transition to psychosis [10, 11]. [^18^F]DOPA elevations in the ventral striatum of schizophrenia patients and of at-risk individuals have also been reported [12, 13]. In parallel, resting-state functional magnetic resonance imaging (fMRI) studies have identified reduced functional connectivity between the dorsal striatum, thalamus, and dlPFC in first episode psychosis (FEP) patients, their unaffected first-degree relatives, at-risk individuals, chronic unmedicated patients, and healthy people with psychosis-like experiences [2–5, 14]. Increased functional connectivity between the ventral striatum, limbic regions, anterior cingulate cortex, ventromedial prefrontal and orbitofrontal cortices has also been found in these groups [2, 15–17]. Correlations between striatal [^18^F]DOPA activation of prefrontal and medial temporal areas have been reported in early and chronic stages of psychotic illness [18–20].

Functional connectivity quantifies statistical dependencies between regional activity and does not distinguish causal interactions. It is therefore unclear whether FST dysfunction in psychosis arises from altered bottom-up signaling or disrupted top-down regulation of subcortical systems. Evidence for a primary bottom-up pathology comes from reports of aberrant molecular function, activity, and functional connectivity of the midbrain across at-risk and schizophrenia groups [21–23]. Others have proposed, largely based on ex-vivo and preclinical findings, that subcortical changes are secondary to deficient top-down control of midbrain neurons, which is caused by GABA/glutamate imbalance in the cortex or hippocampus [9, 24, 25]. In-vivo imaging findings have been mixed, with reports of increased, decreased, or no changes in prefrontal excitatory or inhibitory metabolites in FEP [26–30] and people with established schizophrenia [27, 28, 31, 32].

Top-down and bottom-up interactions between distinct elements of FST systems can be disentangled through models of effective connectivity. Effective connectivity refers to the causal influence that one neural system exerts over another [33] and can be estimated from fMRI data using dynamic causal modelling (DCM). DCM uses a Bayesian framework for developing a generative model of directed, or causal, influences between regions comprising a distributed neural system [33]. Here, we used spectral DCM [34] to characterize disruptions of dorsal and ventral FST effective connectivity in antipsychotic-naïve FEP patients and established schizophrenia (SCZ) patients using resting-state fMRI data. In an independent healthy cohort who underwent concurrent fMRI and [^18^F]DOPA PET we then identified the specific FST connections that are associated with dorsal and ventral striatal [^18^F]DOPA levels. This approach allowed us to cross-sectionally map effective dysconnectivity of FST circuits across different illness stages and to identify putative directed influences associated with striatal dopamine synthesis capacity.

## Methods

### Participants

Detailed information about the participants, recruitment, data exclusion, and symptom measures in Supplementary Information. Briefly, we examined three independent cohorts: (1) a FEP cohort comprising antipsychotic-naïve patients within six months of psychosis onset and healthy controls [35]; (2) a SCZ cohort comprising schizophrenia patients and healthy controls obtained through the UCLA Consortium for Neuropsychiatric Phenomics open dataset [36]; and (3) an [^18^F]DOPA cohort comprising healthy participants recruited from the community who underwent concurrent PET/fMRI scans. Separate analyses were also performed for a subgroup of the FEP cohort with a schizophrenia diagnosis (FEP-SCZ). Group numbers and demographic details are provided in Table 1.

**Table 1.**
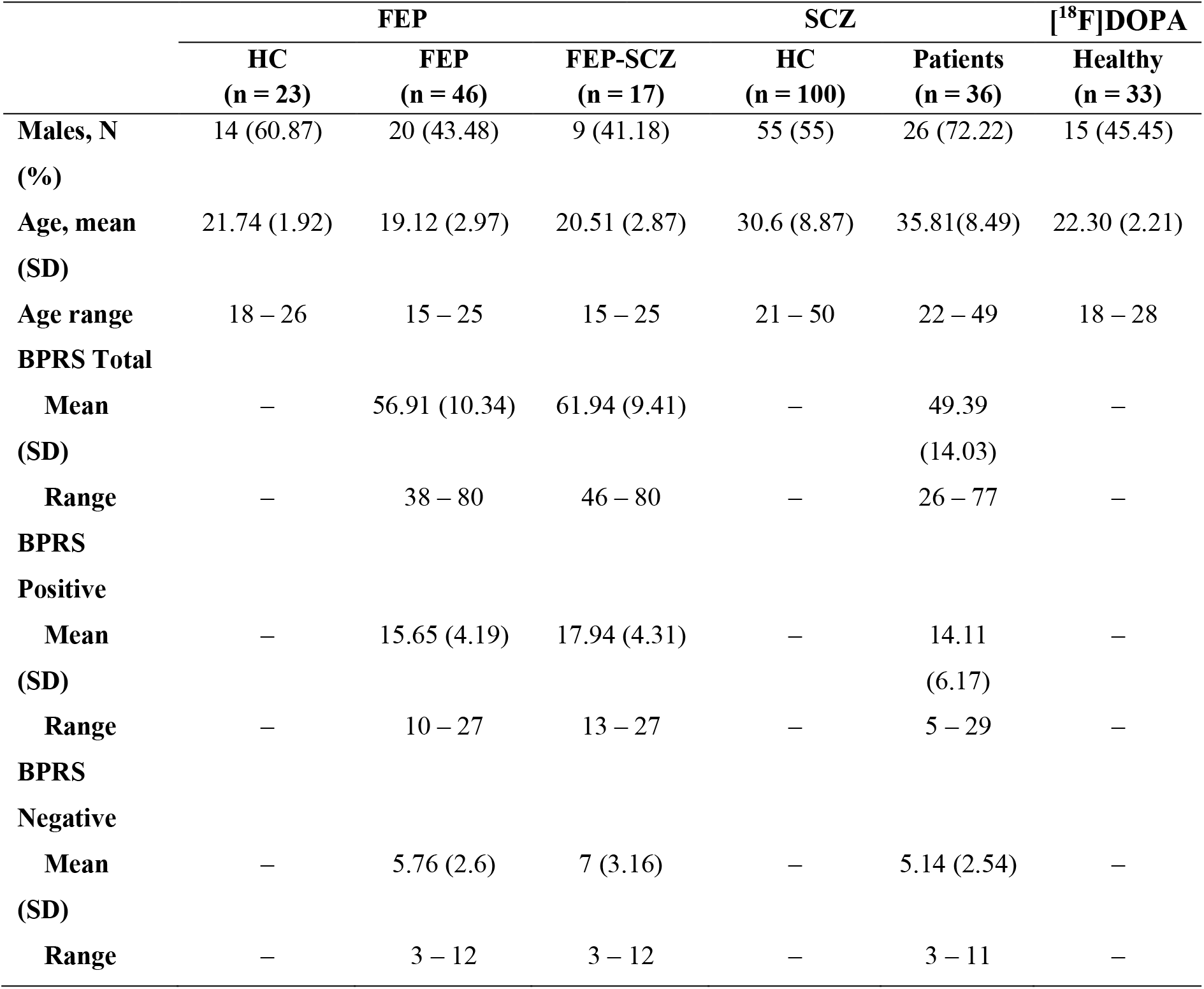
Participants Descriptive Statistics.

|  | FEP |  |  | SCZ |  | [<sup>18</sup>F]DOPA |
| --- | --- | --- | --- | --- | --- | --- |
|  | HC<br>(n = 23) | FEP<br>(n = 46) | FEP-SCZ<br>(n = 17) | HC<br>(n = 100) | Patients<br>(n = 36) | Healthy<br>(n = 33) |
| **Males, N** | 14 (60.87) | 20 (43.48) | 9 (41.18) | 55 (55) | 26 (72.22) | 15 (45.45) |
| **(%)** |  |  |  |  |  |  |
| **Age, mean** | 21.74 (1.92) | 19.12 (2.97) | 20.51 (2.87) | 30.6 (8.87) | 35.81(8.49) | 22.30 (2.21) |
| **(SD)** |  |  |  |  |  |  |
| **Age range** | 18 – 26 | 15 – 25 | 15 – 25 | 21 – 50 | 22 – 49 | 18 – 28 |
| **BPRS Total** |  |  |  |  |  |  |
| **Mean** | – | 56.91 (10.34) | 61.94 (9.41) | – | 49.39 | – |
| **(SD)** |  |  |  |  | (14.03) |  |
| **Range** | – | 38 – 80 | 46 – 80 | – | 26 – 77 | – |
| **BPRS** |  |  |  |  |  |  |
| **Positive** |  |  |  |  |  |  |
| **Mean** | – | 15.65 (4.19) | 17.94 (4.31) | – | 14.11 | – |
| **(SD)** |  |  |  |  | (6.17) |  |
| **Range** | – | 10 – 27 | 13 – 27 | – | 5 – 29 | – |
| **BPRS** |  |  |  |  |  |  |
| **Negative** |  |  |  |  |  |  |
| **Mean** | – | 5.76 (2.6) | 7 (3.16) | – | 5.14 (2.54) | – |
| **(SD)** |  |  |  |  |  |  |
| **Range** | – | 3 – 12 | 3 – 12 | – | 3 – 11 | – |

Positive symptoms were assessed in patients using the positive subscale of Brief Psychiatric Rating Scale (BPRS) version 4 [37]. Secondary analyses also considered negative symptoms assessed with the same instrument.

### MRI/PET processing and analysis

Echo-planar images (EPI) were preprocessed using FMRIPREP (version 1.1.1) [38], with individuals subjected to rigorous denoising and quality control for motion artifacts, as per past work [39]. Acquisition and processing details are in Supplementary Methods.

PET data in the [^18^F]DOPA group were acquired on a MR-PET Siemens Biograph scanner, following bolus injection of approximately 150 MBq of [^18^F]DOPA. Patlak graphical analysis was used to quantify [^18^F]DOPA influx rate constants (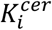 values) for dorsal and ventral striatal regions-of-interest (ROIs) [40] (Figure S1) relative to the cerebellum [41] in each participant’s anatomical space. We only analyzed the left hemisphere consistent with our DCM (see below). Further details are in Supplementary Methods.

### Dynamic causal modeling

#### Model space selection

Regions of interest (ROIs) spanning the dorsal and ventral FST systems and the midbrain were selected using stereotactic coordinates of past findings or of peak signals identified using functional connectivity, as outlined in Supplementary Methods. We modelled 47 biologically plausible connections between eight ROIs (Figure 1). Due to the coarse resolution of the scans, we combined ventral tegmental area and substantia nigra (VTA/SN) into one midbrain ROI. We focused on the left hemisphere given prior evidence of more consistent functional connectivity effects in left-lateralized FSTs [2, 3].

**Figure 1.**
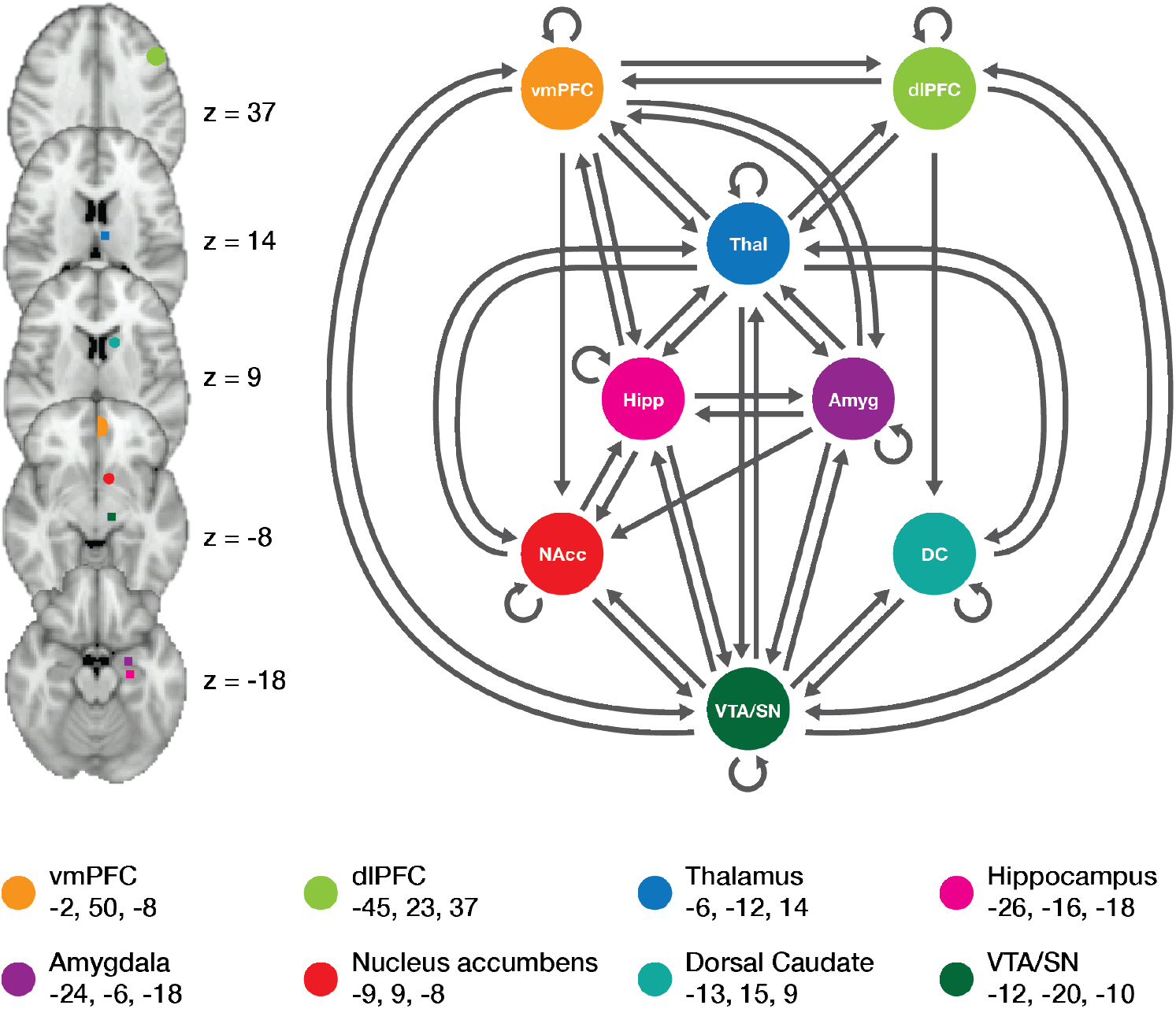
Parent model space of fronto-striato-thalamic systems encompassing dorsal and ventral circuits. Left shows anatomical locations of ROIs on axial slices. Right shows the parent model, including 47 biologically plausible connections including self-connections (circular arrows) for a network comprising eight regions. Centroids of ROIs in MNI coordinates (x, y z) are presented in the bottom panel. VTA/SN: ventral tegmental area/substantia nigra (i.e., the midbrain).

#### Model estimation

We modelled effective connectivity in the spectral domain by fitting a complex cross spectral density using a generative model, parametrized by a power-law model of endogenous fluctuations, implemented in SPM12 (DCM 12; revision 7487) [34]. Details are provided in Supplementary Methods. Briefly, subject-specific first-level analyses were used to estimate directed (causal) influences between regions (in Hz), and the (inhibitory) recurrent or self-connectivity (i.e., self-inhibition) within each region. Following first-level model inversion, we excluded subjects with <75% variance explained by DCM for subsequent analyses (details in Supplementary Methods).

Subject-specific connectivity parameters were then passed to a group-level general linear model (GLM) implemented in the parametric empirical Bayes (PEB) framework to estimate between-subject variability [42]. PEB models were run separately for each cohort. FEP and SCZ patients were compared to their respective control groups. Symptom correlates in patients were modelled separately for each group, with both positive and negative symptoms included as covariates. Associations with [^18^F]DOPA were modelled in healthy individuals only, with dorsal and ventral striatal 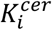 values as covariates. Age, sex, and mean framewise displacement as a measure of in-scanner motion [43] were used as nuisance covariates for all models. Scanner site was also used as a covariate in the SCZ group. We only report effects with a posterior probability threshold above 0.95. PEB is a multivariate (Bayesian) GLM in which we fit all model parameters at once; hence, no correction for multiple comparisons is required. A typical effect size for effective connectivity between regions is 0.1 Hz [34]. Self-connections are inhibitory to reflect activity decay, with a negative sign indicating reduced self-inhibition (i.e., disinhibition, or slower activity decay), and a positive sign indicating increased self-inhibition (i.e., quicker activity decay). Technical details are outlined in Supplementary Methods.

## Results

### Group differences in effective connectivity

#### FEP

Patients showed reduced self-inhibition of VTA/SN, increased self-inhibition of dlPFC, greater inhibitory influence of the thalamus on NAcc, and an excitatory influence of the amygdala on NAcc relative to controls (Figure 2A; Table S1). The subset of FEP-SCZ patients (Figure S2) showed similar changes, but did not show increased dlPFC self-inhibition. FEP-SCZ patients additionally demonstrated a disinhibition of the amygdala, increased excitatory influence of VTA/SN on NAcc, and increased inhibitory influence of VTA/SN and NAcc on hippocampus. Connections between cortical and subcortical areas were not implicated as dysfunctional in FEP patients.

**Figure 2.**
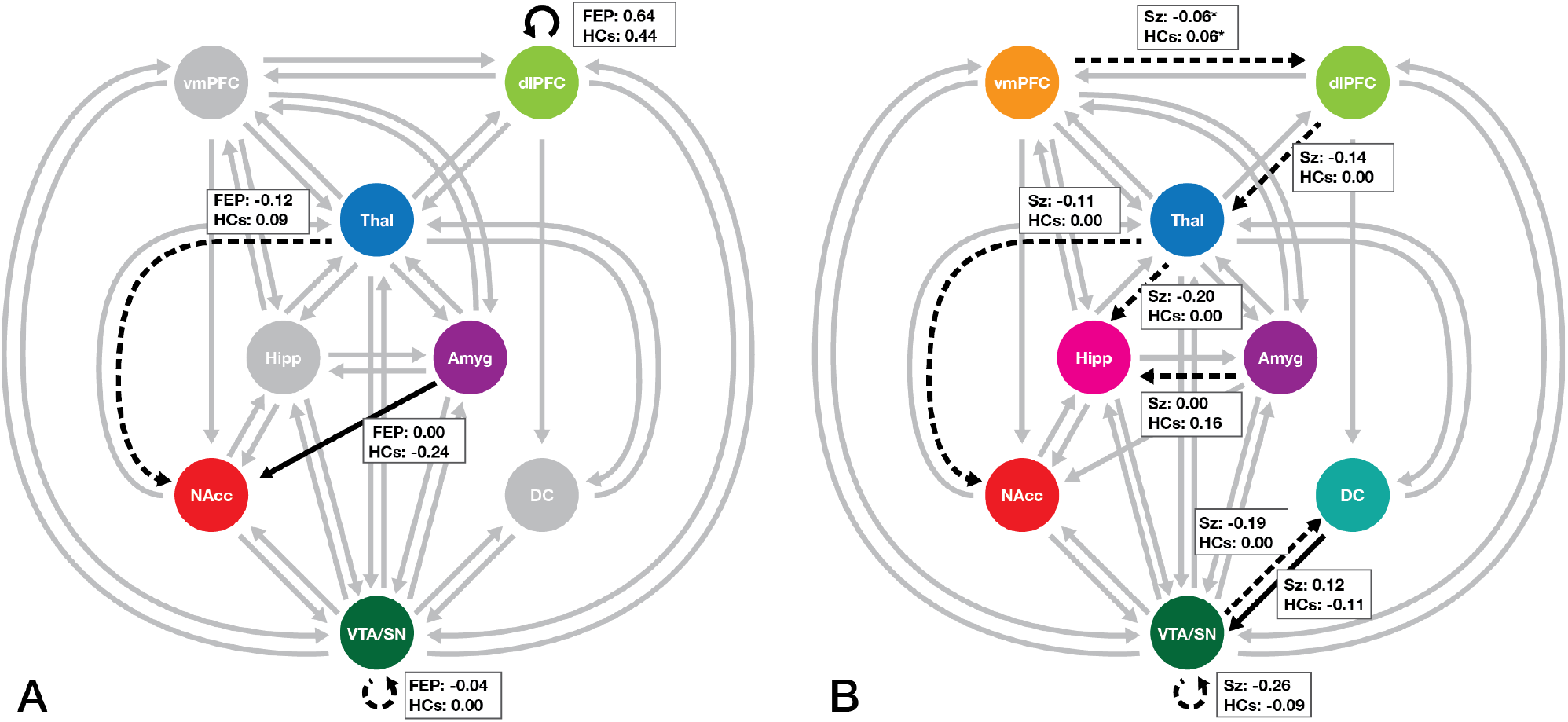
Group differences in fronto-striato-thalamic effective connectivity identified in the FEP (n = 46) and SCZ (n = 36) patients relative to their respective control groups. Differences in effective connectivity between FEP patients and healthy controls are shown in panel A, and patients with established schizophrenia and healthy controls are shown in panel B. Boxes show mean connectivity values in each group. For connections between regions, dashed arrows represent connections for which patients show a stronger inhibitory influence compared to controls; solid arrows represent connections for which patients show stronger excitatory influence compared to controls. For self-connections, dashed arrows represent reduced inhibition and solid arrows indicate increased inhibition in patients compared to controls. Gray arrows represent modeled connections that were not significantly different from the prior. Inter-regional connectivity parameters are in Hz. Self-connection values are in negative log scale. Connections were thresholded at Pp > 0.95 which represents strong evidence. DC: dorsal caudate; NAcc: nucleus accumbens: VTA/SN: ventral tegmental area/substantia nigra (midbrain). * denotes mean connections that were derived from the DCM PEB routine as these connections were removed in the subsequent Bayesian model averaging routine. * denotes mean connections that were derived from the DCM PEB routine as these connections were removed in the subsequent Bayesian model averaging routine.

#### SCZ

Similar to FEP, SCZ patients showed disinhibition of VTA/SN and greater inhibitory influence of thalamus on NAcc relative to controls (Figure 2B; Table S1). SCZ patients also showed increased inhibitory influence of vmPFC on dlPFC, of dlPFC on thalamus, and of thalamus and amygdala on hippocampus. FEP-SCZ patients and SCZ patients both showed altered bottom-up connectivity of the VTA/SN. However, where the disruptions primarily affected the limbic circuit in FEP-SCZ, they affected the dorsal system in SCZ, with increased inhibitory influence on the dorsal caudate (DC) and increased excitatory influence of DC on VTA/SN.

### Associations with positive symptoms

#### FEP

Greater positive symptom severity was predominantly associated with subcortical connectivity (Figure 3, Table S2). Specifically, more severe symptoms were associated with a stronger influence of VTA/SN on NAcc and amygdala, of NAcc and hippocampus on VTA/SN. More severe symptoms were also associated with a weaker influence of VTA/SN on DC and hippocampus, and with reduced self-inhibition of the amygdala. In cortex, higher positive symptom ratings were associated with reduced influence of dlPFC on vmPFC and reduced self-inhibition of vmPFC.

**Figure 3.**
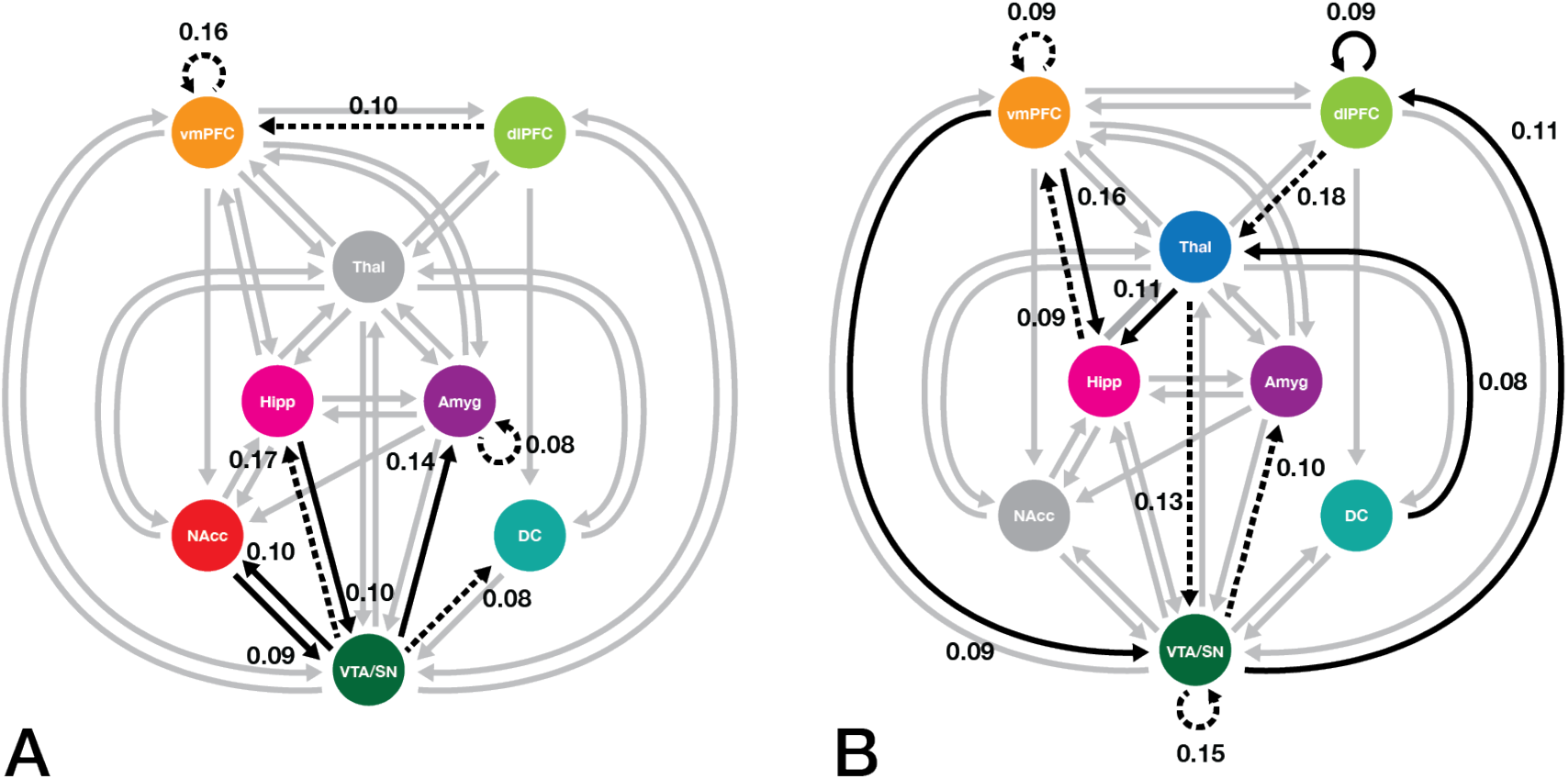
Associations between positive symptoms and fronto-striato-thalamic effective connectivity parameters in the FEP (n = 46) and SCZ (n = 36) patients. Panels from left to right depict the results of PEB models mapping associations between FST effective connectivity and positive symptoms in (A) FEP and (B) SCZ patients. For between-region connections, solid arrows denote positive associations and dashed arrows depict negative associations between effective connectivity parameters and symptoms. For self-connections, solid arrows represent positive associations between symptom severity and self-inhibition, whereas dashed arrows denote negative associations such as more severe symptoms were associated with reduced inhibition. Gray arrows show modelled associations that were not (significantly) different from the prior. Connections were thresholded at Pp > 0.95. DC: dorsal caudate; NAcc: nucleus accumbens: VTA/SN: ventral tegmental area/substantia nigra (midbrain).

The FEP-SCZ subgroup (Figure S3, Table S2) showed similar associations with vmPFC self-inhibition and subcortical connectivity, although associations with hippocampal to VTA/SN connectivity and amygdala self-inhibition were absent. Positive symptom severity was additionally associated with a weaker influence of NAcc on hippocampus, of hippocampus on amygdala, of DC on thalamus, and reduced dlPFC self-inhibition, coupled with a stronger influence of dlPFC on thalamus and increased thalamic self-inhibition.

#### SCZ

Both cortical-subcortical and subcortical-cortical influences were associated with positive symptoms in SCZ patients (Figure 3, Table S2). Specifically, more severe positive symptoms were associated with stronger top-down influence of vmPFC on VTA/SN and hipppocampus, stronger bottom-up influence of VTA/SN on dlPFC, stronger influence of DC on thalamus, of thalamus on hippocampus, and greater dlPFC self-inhibition. Higher positive symptom ratings were also associated with reduced influence of dlPFC on thalamus, thalamus on VTA/SN, VTA/SN on amygdala, hippocampus on vmPFC, and vmPFC and VTA/SN self-inhibition. Some common connections were implicated in both FEP-SCZ and SCZ patients (i.e., VTA/SN–amygdala, DC– thalamus, dlPFC–thalamus, and dlPFC self-inhibition), albeit with opposing polarity, suggesting that the link between positive symptoms and altered FST effective connectivity varies across illness stages.

### Associations with negative symptoms

Associations with negative symptom for FEP, FEP-SCZ, and SCZ patients are presented in Figure S4 and Table S2. Associations between symptom severity and amygdala self-inhibition were consistent, albeit with reversed polarity, across the last two groups. Top-down cortical to subcortical connections were associated with negative symptom severity in FEP-SCZ patients, whereas SCZ patients showed associations with bottom-up subcortical to cortical connections. Similar to positive symptoms, negative symptom associations in the larger group of FEP patients mainly implicated subcortical connections.

### Associations with striatal dopamine synthesis capacity

#### Dorsal striatum

Significant associations with dorsal striatal [^18^F]DOPA in healthy individuals largely implicated the thalamus (Figure 4; Table S3). Specifically, higher [^18^F]DOPA was associated with stronger thalamic self-inhibition, stronger thalamic influence over the VTA/SN and DC, and amygdala over thalamus, in addition to a weaker influence of DC on thalamus. Increased bottom-up influence of the amygdala on vmPFC was also associated with higher dorsal striatal [^18^F]DOPA.

**Figure 4.**
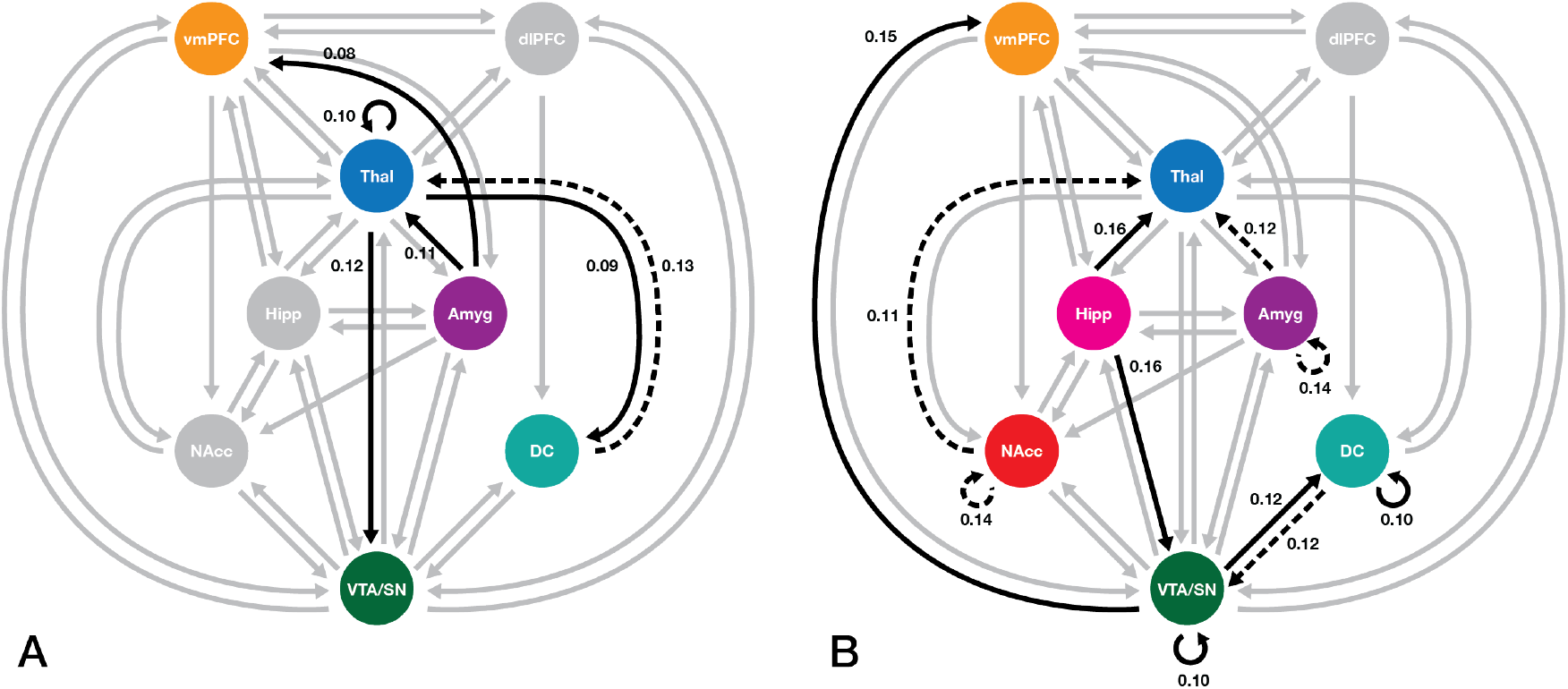
Associations between fronto-striato-thalamic connectivity and striatal dopamine synthesis capacity. Panel A depicts connections associated with dopamine synthesis in the dorsal circuit. Panel B illustrates associations with dopamine synthesis in the ventral circuit. Solid arrows: positive associations between effective connectivity parameters and dopamine synthesis; dashed arrows: negative associations between effective connectivity parameters and dopamine synthesis; gray arrows: modelled connections that were not (significantly) different from the prior. For self-connections, solid arrows represent positive associations between striatal dopamine synthesis and self-inhibition, whereas dashed arrows denote negative associations between striatal dopamine synthesis and self-inhibition. Connections were thresholded at Pp > 0.95. DC: dorsal caudate; NAcc: nucleus accumbens: VTA/SN: ventral tegmental area/substantia nigra (midbrain).

#### Ventral striatum

Ventral striatal [^18^F]DOPA was associated with a distributed set of extrinsic effective connections centered on midbrain and thalamus (Figure 4; Table S3). Specifically, higher [^18^F]DOPA was associated with weaker self-inhibition of NAcc and amygdala, weaker influence of these two regions on the thalamus, and stronger influence of hippocampus on thalamus. Ventral striatal [^18^F]DOPA was also associated with stronger self-inhibition of the VTA/SN and influence of this region on the vmPFC and DC, respectively weaker and stronger influence of DC and hippocampus on VTA/SN, and stronger DC self-inhibition.

## Discussion

FST dysfunction has been identified in psychotic illnesses [2–5, 14–17], but a characterization of how causal influences within these circuits are altered at distinct illness stages has been lacking. Using spectral DCM, we mapped the effective connectivity of dorsal and ventral FST circuits in FEP and established SCZ. Both clinical groups showed consistent disinhibition of VTA/SN and a stronger top-down inhibitory influence of the thalamus on NAcc. Altered top-down connectivity from the cortex to subcortex was only identified in established illness. Positive symptom severity was associated with self-disinhibition of the VTA/SN in both illness stages. Additional associations were otherwise largely confined to subcortical connectivity in FEP and a distributed set of cortical and subcortical connections in SCZ patients. Concurrent PET-MRI in healthy individuals revealed distinct sets of FST connections associated with dorsal and ventral dopamine synthesis capacity, with DC and striatothalamic connectivity associated with both striatal dopamine synthesis and positive symptom severity. Our findings indicate that midbrain dysfunction and subcortical dysconnectivity is prominent in early illness stages, that cortical dysfunction becomes more salient in established illness, and that striatothalamic and nigrostriatal connectivity are related to both striatal dopamine synthesis capacity and positive symptom severity in patients.

### Effective dysconnectivity of fronto-striato-thalamic circuitry

We observed VTA/SN disinhibition across both illness stages, suggesting that it represents a core feature of FST pathology in psychosis. Midbrain disinhibition is consistent with elevated striatal dopamine synthesis [10]. Given that we found no evidence of disrupted top-down influences on VTA/SN in FEP, our findings thus suggest that prior reports of elevated striatal dopamine in [^18^F]DOPA and other PET studies across different stages of psychosis may be linked to intrinsic dysregulation of the midbrain [25, 44].

Both FEP and SCZ groups also showed increased inhibitory influence of thalamus over NAcc. Excitatory thalamostriatal projections provide feedback for the striatum to maintain bottom-up signaling to thalamus and cortex in support of specific actions or behaviors [45]. A greater inhibitory influence of thalamus on NAcc in both FEP and SCZ thus implies a dysregulation of the thalamostriatal feedback pathway. Although this dysregulation was not directly tied to VTA/SN function in our findings, animal models suggest that NAcc dysregulation can disinhibit the midbrain in young adult rats [25, 46].

SCZ patients showed increased inhibitory influence of VTA/SN over DC, which should yield a net disinhibition of striatal activity given that ∼95% of striatal neurons are GABAergic [47]. Striatal disinhibition may disrupt the capacity of this area to filter information through the FST circuits [9]. SCZ patients also showed increased top-down excitatory influence of DC over VTA/SN, which may reflect a compensatory response to regulate the disinhibitory influence of midbrain over the DC.

Given that VTA/SN disinhibition was also observed in FEP, our findings suggest that an early bottom-up pathology of the midbrain may evolve to affect striatal function and potentially dysregulate feedback loops within the dorsal FST with illness progression. However, we cannot rule out an effect of medication in the SCZ group and longitudinal data are required to test this hypothesis.

In FEP, cortical dysfunction was limited to increased dlPFC self-inhibition. Post-mortem work suggests robust changes in GABAergic neuron function in patients with established schizophrenia [48], but in vivo studies of FEP patients have been inconsistent, with reports of lower, higher, or no differences in prefrontal GABA levels [29, 30, 49, 50]. Our findings thus suggest that while DLPFC dysfunction is evident early in psychosis, it does not to play a prominent role in broader FST dysfunction.

Taken together, our DCM analysis indicates that the early phase of psychosis is associated with prominent dysfunction of subcortical systems, with alterations in cortico-subcortical connectivity emerging in later illness stages. Intrinsic dysfunction of the VTA/SN and dlPFC, and altered thalamic regulation over the NAcc, appear to be stable features of the illness.

### FST circuit connectivity and symptom severity

A primary role for VTA/SN dysregulation in psychosis is supported by our observations of consistent correlations between the connectivity of this region and positive symptom severity across illness stages [23, 51–53]. However, the specific connections implicated varied across the patient groups. In the full FEP cohort, greater positive symptom severity was associated with connectivity between VTA/SN and limbic subcortical regions, which accords with a role for hippocampal–midbrain dysregulation identified in rodent models of psychosis [25]. Disinhibition of the vmPFC, another component of the limbic FST system, was negatively associated with positive symptom severity across the FEP, FEP-SCZ, and SCZ patients [54].

Reduced influence of VTA/SN on DC was also associated with greater positive symptom severity in both FEP-SCZ and SCZ patients. When taken with evidence of robust [^18^F]DOPA elevations in the DC in early psychosis [10, 11, 55], our findings of intrinsic dysfunction of the midbrain across illness stages [see also 23–25], and anatomical studies indicating that midbrain afferents to the dorsal striatum primarily originate in the SN [8], and our results identifying a close link between nigrostriatal signaling and the expression of psychotic symptoms. Several additional FST elements were consistently associated with positive symptom severity in both SCZ and FEP-SCZ patients, although the polarity of associations was often reversed between FEP and SCZ groups, which may reflect differences in the neural correlates of psychotic symptoms at different illness stages, an effect of medication in the SCZ group, or a combination of both.

There was less consistency across the cohorts with respect to negative symptom associations, although the dlPFC and limbic regions were generally implicated, consistent with evidence of prefrontal and limbic involvement in this symptom dimension [56–58]. Cortical-subcortical connections were implicated in FEP-SCZ and SCZ patients, with the former mainly demonstrating associations with top-down influences and the latter group showing associations with bottom-up influences.

### FST circuit connectivity and striatal dopamine synthesis capacity

Dorsal and ventral striatal [^18^F]DOPA levels were associated with effective connectivity of distinct FST circuit elements. Most associations with dorsal striatum [^18^F]DOPA were limited to the dorsal FST circuit and involved the thalamus. Two thalamic connections––an afferent input from DC and the thalamic self-connection––were also associated with positive symptom severity in FEP-SCZ patients. The modulation of cortical and thalamic glutamatergic signals by striatal dopamine controls striatothalamic filtering and information flow to the cortex, which is thought to play a central role in the pathogenesis of psychosis [9].

Bidirectional connectivity between the VTA/SN and DC was associated with dorsal striatal [^18^F]DOPA, identified as dysfunctional in SCZ patients, and was correlated with positive symptom severity in both SCZ and FEP-SCZ, suggesting a close link between nigrostriatal signaling, striatal dopamine synthesis capacity, and positive symptom severity in patients. Notably, positive symptom severity in FEP was associated with a weaker influence of VTA/SN on DC whereas higher dorsal striatal [^18^F]DOPA was associated with a stronger afferent influence of VTA/SN. Since patients are known to show elevated [^18^F]DOPA in the dorsal striatum [10, 11], one might expect a negative association between striatal [^18^F]DOPA and VTA/SN-DC connectivity. One potential explanation is that elevated levels in dopamine lead to compensatory changes in circuit function that affect the optimal balance of signaling between different regions, thus resulting in different circuit mechanisms that regulate dopamine levels in the brains of psychotic and non-psychotic individuals. Concurrent PET-fMRI in both patients and controls would help to test this possibility.

Connections associated with ventral striatal [^18^F]DOPA were distributed across the ventral and dorsal systems. NAcc projections are widely distributed in the midbrain and drive dorsal striatal dopamine levels [8]. Accordingly, we found that ventral striatal [^18^F]DOPA was associated with VTA/SN–DC connectivity and inhibition within both regions. VTA/SN–DC connectivity was also disrupted in SCZ patients and associated with positive symptoms in FEP, suggesting a link between the NAcc’s regulation of midbrain activity and dorsal circuit dysfunction in patients [59].

### Limitations and future directions

We used independent samples to cross-sectionally characterize effective dysconnectivity across distinct illness stages. This approach offers a test of consistency across cohorts, but inferences about the progression of circuit dysfunction must be confirmed longitudinally, especially given the differences in medication exposure, sample size, and symptom severity across the three cohorts.

Our PEB analysis of the PET data identifies associations with, but not causal influences on, striatal [^18^F]DOPA. Further investigation of patients would be required to identify precisely how effective dysconnectivity of FST systems leads to dopamine dysregulation in patients.

We restricted our analysis to the left hemisphere because it has been more frequently as relevant for psychosis by prior work [2, 3]. This focus facilitates efficient estimation of DCMs but may miss influences of wider brain regions connected with the striatum. Recent improvements in the scalability of DCM [60] may be used to derive a more comprehensive picture..

## Conclusions

Our analysis characterized FST effective dysconnectivity in FEP and established schizophrenia. Our findings indicate that subcortical dysfunction features prominently in early illness stages, with cortical abnormalities more apparent later in the illness. In early psychosis, positive symptoms are associated with midbrain connectivity, suggesting that aberrant bottom-up signals emanating from the midbrain are a key feature in the pathogenesis of psychotic symptoms. Nigrostriatal and striatothalamic connectivity are closely linked to striatal dopamine levels while also being tied to symptom expression in patients. Together, our findings suggest a prominent role for subcortical systems in driving FST dysfunction in psychosis.

## Data Availability

Data for FEP and FDOPA participants are available upon request. Data for schizophrenia patients and their healthy controls are open source and are available at https://openneuro.org/datasets/ds000030/versions/00016

## Disclosures

The authors declare no financial relationships with commercial interests

## Acknowledgments

This work was supported by the National Health and Medical Research Council (NHMRC) (ID: 1050504), Australian Research Council (ID: FT130100589) and the Charles and Sylvia Viertel Charitable Foundation.

Pantelis was supported by a NHMRC Senior Principal Research Fellowship (ID: 1105825). In the past 5 years, Pantelis served on an advisory board for Lundbeck, Australia Pty Ltd. He has received honoraria for talks presented at educational meetings organized by Lundbeck. We thank Hannes Almgren and Ian Harding for advice on data analysis. The authors acknowledge the facilities and scientific and technical assistance of the National Imaging Facility, a National Collaborative Research Infrastructure Strategy (NCRIS) capability.

## Notes

### Competing Interest Statement

The authors have declared no competing interest.

### Author Declarations

Melbourne Health Human Research Ethics Committee (IDs: 2007.616 and 1443066.1)

