## Supplementary Information for "Effective Connectivity and Dopaminergic Function of Fronto-Striato-Thalamic Circuitry in First-Episode Psychosis, Established Schizophrenia, and Healthy Controls"

**Appendix.** Supplementary Methods and Materials

**Table S1.** Summary of Group Differences in Fronto-striato-thalamic Effective Connectivity in FEP (n = 46), FEP-SCZ (n = 17), and SCZ (n = 36) Patients Respective to Healthy Controls

**Table S2.** Summary of Connections Associated with Severity of Clinical Symptoms in FEP (n = 46), FEP-SCZ (n = 17), and SCZ (n = 26) Patients

**Table S3.** Summary of Connections Associated with PLEs (n= 26) and Striatal Dopamine Synthesis (n = 33) in the Healthy Cohort

**Figure S1.** Parcellation of Dorsal and Ventral Striatal Regions of Interest in PET Quantification Presented on the MNI 2mm Template

**Figure S2.** Group Differences in Effective Connectivity Parameters Between FEP-SCZ Patients (N = 17) and Healthy Controls

**Figure S3.** Effective Connectivity Associated with Positive Symptoms in FEP-SCZ Patients (N = 17)

**Figure S4.** Associations Between Severity of Negative Symptomatology and Effective Connectivity Parameters Across All Cohorts

**Appendix. Supplementary Methods and Materials**

**Participants**

**FEP.** FEP was defined as fulfilling Structured Clinical Interview for DSM-5 (SCID-5) criteria for a psychotic disorder. Patients with substance-induced psychosis were excluded in the present study. FEP diagnoses are in Table S2. Healthy controls had no history of psychiatric or neurological illness, as determined by self-reports, SCID, and the Comprehensive Assessment of At-Risk Mental States. All participants provided informed consent. For FEP patients, additional strict inclusion criteria include comprehension of English language, no contraindication to MRI scanning, less than six months of duration of untreated psychosis, living in stable accommodation, low risk to self or others (score of < 5 on the Brief Psychiatric Rating Scale version 4 (BPRS-4) Suicidality and Hostility subscales), and minimal previous exposure to antipsychotic medication (less than 7 days of use or lifetime 1750mg chlorpromazine equivalent exposure). For healthy controls, additional exclusion criteria included history of psychiatric or neurological illness in first-degree relatives and current use of psychotropic medications. Only baseline data were used in the present study. We also analyzed a subsample of FEP patients with schizophrenia diagnoses (FEP-SCZ).

The initial sample size for this group was 61 patients and 27 controls. We excluded 5 patients with poor imaging data, 1 patient with high motion, 3 patients with DCM explaining <75% of the signal variance, and 6 patients with substance induced psychosis. This brought the final sample of patients included in the study to 46. A total of 17 FEP patients had a diagnosis of schizophrenia spectrum disorder (8 schizophrenia, 8 schizophreniform disorder and 1 schizoaffective disorders), making up a subset of FEP-SCZ patients. Other FEP patients were diagnosed with delusional disorder (n = 5), major depressive disorder with psychotic features (n = 10), psychotic disorder not otherwise specified (n = 13), and 1 patient had a missing diagnosis. For the control group, we excluded 3 individuals with high motion (criteria outlined below), and 1 with DCM explaining <75% of the signal variance, resulting in a final sample of 23.

**Established schizophrenia (SCZ).** Data were obtained through the UCLA Consortium for Neuropsychiatric Phenomics open dataset (Poldrack et al., 2016). The initial sample consisted of 121 right-handed healthy controls and 51 schizophrenia patients. Healthy controls were excluded if they had a history of mental illness. Schizophrenia patients were excluded if they had comorbidity with either bipolar disorder or ADHD. Diagnoses were based on the DSM-IV and SCID-1.

We excluded 1 patient with poor imaging data, 9 with high motion, and 5 with DCM explaining <75% of the signal variance, resulting in a final sample of 36 patients. We excluded 6 controls with high motion and 15 with DCM explaining <75% of the signal variance, resulting in a final sample of 100 controls.

**[$^{18}$F]DOPA.** A total of 52 healthy participants were recruited from the community through online advertisements. Each participant provided informed consent. Exclusion criteria included current or history of psychiatric or neurological illnesses, significant medical history, intellectual disability, and first-degree relative with a mental illness. Participants underwent simultaneous resting-state fMRI and [$^{18}$F]DOPA PET.

Eight individuals did not complete the scanning protocol. We also excluded 3 participants with high motion and 5 participants with DCM explaining <75% of the signal variance. A total of 3 participants had unusable PET scans, bringing the final sample to 33 with fMRI and PET data.

**Symptom measures**

The Brief Psychiatric Rating Scale (BPRS) positive and negative subscales were used to respectively measure positive and negative symptoms in our clinical cohorts. The subscales were derived from a five factor solution outlined in Dazzi et al., 2016. The positive symptom subscale consists of five items: unusual thought content, suspiciousness, hallucinations, grandiosity, and bizarre behavior. The negative symptom subscale consists of three items: blunted affect, emotional withdrawal, and motor retardation. Each item is ranked on a Likert-like scale between 1 to 7, with a score of 1 denoting the absence of a measured symptom.

**Neuroimaging acquisition**

**FEP.** Whole-brain T2*-weighed echo-planar images (EPIs) and anatomical T1-weighted (T1w) scans were acquired for each participant using a 3T Siemens Trio Tim scanner, equipped with a 32-channel head coil, located at the Royal Children's Hospital in Melbourne, Australia. Participants were instructed to lie still in the scanner while maintaining wakefulness with eyes closed. A total of 234 functional volumes with 37 slices each were acquired using the following parameters: repetition time = 2000ms; echo time = 32ms; flip angle = 90°; field of view = 210mm; slice thickness of 3.5 mm, and 3.3 x 3.3 x 3.55 mm voxels. A total of 176 slices were acquired for each participant's T1-weighted image using an interleaved acquisition using the following parameters: TR = 2.3s; TE = 2.98ms; flip angle of 9°; FOV of 256mm; voxel size of 1.1 x 1.1. x 1.2 mm.

**SCZ.** The CNP dataset (Poldrack et al., 2016) was acquired on one of two Siemens Trio 3T scanners located at the Ahmanson-Lovelace Brain Mapping Center and the Staglin Center for Cognitive Neuroscience at UCLA. Details of the resting-state EPI scan are TR = 2s, TE = 30ms , flip angle = 90°, 4mm slice thickness, 152 volumes with 34 slices each. Participants were instructed to keep their eyes open. Details of the T1 scan are TR = 1.9s, TE = 2.26ms, flip angle of 90°, 176 slices with $1mm^3$ voxels.

**[$^{18}$F]DOPA.** Data were acquired using a 3T Siemens Magnetom Biograph simultaneous MR-PET scanner equipped with a 20-channel head and neck coil at Monash Biomedical Imaging, Melbourne, Australia. Resting-state whole-brain T2*-weighed echo-planar image (EPIs) was also acquired for each subject using an interleaved acquisition with the following parameters: TR = 2.89s; TE = 30ms; 152 volumes with 44 slices per volume; flip angle = 90°; FOV = 190mm; slice thickness = 3mm; voxel size of $3mm^3$. A high resolution T1-weighted anatomical image was acquired for each subject using an ascending acquisition (176 slices; TR = 1640ms; TE = 2.34ms; flip angle = 8°; field of view (FOV) = 256mm; slice thickness = 1mm; voxel size = $1mm^3$).

**fMRIPREP preprocessing steps**

fMRI and T1-weighted data were processed in the same way across all three cohorts using FMRIPREP software version 1.1.1 (Esteban et al., 2019). Each T1w scan was corrected for non-uniformity in intensity and subsequently skull stripped. Brain surfaces were reconstructed using FreeSurfer version 6.0.1. Tissue masks were generated using FreeSurfer. Spatial normalization of the skull stripped T1w images to the ICBM 152 Nonlinear Asymmetrical template version 2009c was performed using a nonlinear registration in ANTs version 2.1.0. Similarly, for each participant, tissue masks were registered from the surface space to the MNI template.

EPIs were slice-timed corrected using AFNI version 16.2.07 and realigned to a mean reference image using FSL. EPIs were distortion corrected using fieldmaps (phasediff-based workflow; https://fmriprep.readthedocs.io/en/stable/api/index.html#sdc-phasediff). For participants with missing fieldmaps, a "fieldmap-less" distortion correction was performed by co-registering the functional image to the intensity-inverted T1w image constrained with an EPI distortion atlas (Treiber et al., 2016). Following distortion correction, EPIs were co-registered with their corresponding T1w using boundary-based registration with nine degrees of freedom using the bbregister routine in FreeSurfer. The motion-correcting transformations, field-distortion-correcting warp, EPI-to-T1w transformation, and T1w-to-MNI template warp were concatenated and applied in a single step using ANTs version 2.1.0. In-scanner head motion was defined as excessive according to previously-defined stringent exclusion criteria (Parkes et al., 2018); namely, if any of the following were met: 1) mean framewise displacement (FD) > 0.20mm; (2) sum of suprathreshold FD spikes > 20%; and (3) any FD > 5mm. FD was calculated using the root mean squared volume-to-volume displacement of all voxels, derived from the six head motion parameter (3 translations, 3 rotations) (Parkes et al., 2018). We did not perform global signal regression as DCM incorporates noise parameter estimates that capture observation noise due to scanner and physiological noise (Friston et al., 2014; Razi et al., 2015) and also because it may remove real neuromodulatory fluctuations in neuronal activity, which are of key interest in the present study (for a discussion, see Aquino et al., 2020; Glasser et al., 2018).

**PET acquisition, reconstruction, and attenuation correction**

All participants received carbidopa (150 mg) and entacapone (400 mg) orally 60 minutes before imaging to reduce the formation of radiolabeled metabolites that can cross the blood-brain barrier and thus confound tracer availability in the striatum (Hoffman et al., 1992; Ruottinen et al., 1995). Participants were instructed to lie still in the scanner with eyes closed.

The pseudo-CT attenuation correction method (Baran et al., 2018; Burgos et al., 2014) was used to correct PET images during image reconstruction. Dynamic PET images were reconstructed using the Siemens e7tools software with image volume size 344×344×127(2.09×2.09×2.03mm$^3$). The Ordinary Poisson-Ordered Subset Expectation Maximization (OP-OSEM) algorithm (3 iterations, 21 subsets) was used with the point spread function (PSF) for partial volume correction. A 5-mm FWHM Gaussian filter was applied to each 3D image volume. For correction of subject motion, the list mode dataset was first binned into 95 frames consisting of one 30-second background frame and ninety-four 60-second frames. Dynamic motion was corrected based on an image registration approach (Chen et al., 2019), where each frame was registered to the last frame using rigid-body transformation implemented in the FSL toolbox (Jenkinson et al., 2002). The final reconstructed dynamic PET images were registered to the corresponding T1 MPRAGE MRI image. Patlak graphical analysis was performed using Qmodeling software (López-González et al., 2019; Patlak & Blasberg, 1985). The cerebellum was chosen as a reference region due to its low [$^{18}$F]DOPA uptake (Moore et al., 2003).

**Regions of interest selection for dynamic causal modeling**

**Nucleus accumbens (NAcc) and dorsal caudate (DC).** For the ventral and dorsal striatum, we seeded the NAcc and DC consistent with our previous works (Dandash et al., 2014; Fornito et al., 2013; Sabaroedin et al., 2019) using a functional parcellation of the striatum (Di Martino et al., 2008) that was delineated based on a meta-analysis of striatal activation in fMRI and PET studies (Postuma & Dagher, 2006). The pallidum, which is the primary output structure of the basal ganglia, was omitted to limit model dimensionality. Its effects will thus be captured through indirect (unmodelled) influences in the DCM.

**dlPFC.** To capture variations associated with risk for psychosis as well as disruptions in clinical groups, the dlPFC ROI was selected based on a peak in which functional connectivity with the DC was reduced in the healthy relatives of FEP patients compared to healthy controls (Fornito et al., 2013). This peak overlapped with an area that also showed reduced connectivity in patients.

**vmPFC.** To choose a region that was most relevant to the ventral circuit, we chose a peak in the vmPFC that showed strong functional connectivity with the NAcc in an independent cohort of 353 healthy adults (Sabaroedin et al., 2019).

**Thalamus.** The thalamus coordinate was selected from a peak in which functional connectivity with the dorsal caudate was reduced in ARMS individuals compared to healthy controls (Dandash et al., 2014).

**Hippocampus, amygdala, and midbrain.** ROIs for the three regions were selected using a similar method. Peak coordinates within anatomical masks for the hippocampus and amygdala were selected in a separate group-level functional connectivity analysis of each ROIs connectivity with the whole brain in an independent cohort of 353 individuals (Sabaroedin et al., 2019). For the hippocampus, we chose the anterior region, using of a hippocampal mask provided in the Freesurfer package (Fischl et al., 2002). The anterior hippocampus is a hippocampal region that is most frequently implicated as dysfunctional in schizophrenia (Small et al., 2011), and it was defined as the region having the MNI coordinate of less than y = -22 (Zeidman & Maguire, 2017). Similar to the hippocampus, we used an amygdala mask provided in FreeSurfer. The midbrain mask was delineated based on the functional connectivity of the ventral tegmental (VTA) and substantia nigra (SN) with the whole brain (Murty et al., 2014). Due to the limited spatial resolution of fMRI images, we combined both of the dorsal and ventral midbrain regions and selected a functional connectivity signal peak in the same cohort that was used in the selection of our hippocampal, amygdala, and vmPFC ROIs.

Spherical ROIs were created with a radius of 6mm for cortical regions (i.e., dlPFC and vmPFC) and 3.5mm for each subcortical ROI. For vmPFC and thalamus ROIs with centroids that were close to the anatomical or functional boundary of the region, we used an anatomical mask from the FreeSurfer package when extracting the first eigenvariate for the ROIs to exclude signal from neighboring regions.

**Spectral dynamic causal modelling**

Dynamic causal modelling (DCM) is a Bayesian framework that infers the directed (causal) connectivity among the neuronal systems – referred to as effective connectivity (Friston, 1994). A new DCM for resting state fMRI was recently proposed based upon a deterministic model that generates predicted cross spectra, referred to as spectral DCM. In order to model resting state activity —in the absence of external stimuli—a stochastic component capturing neural fluctuations is included in the model.

Mathematically, we can express the formulation of the stochastic generative model as a set of two equations. First is the neuronal state equation, namely

$$\dot{x}(t) = f(x(t), u(t), \theta) + v(t), \tag{S1}$$

and second is the observation equation, which is a static nonlinear mapping from the hidden physiological states in (1) to the observed BOLD activity, and is written as:

$$y(t) = h(x(t), \varphi) + e(t), \tag{S2}$$

where $\dot{x}(\mathrm{t})$ is the rate of change of the neuronal states $x(t)$, $\theta$ are unknown parameters (i.e., the effective connectivity) and $v(t)$ (resp. $e(t)$) is the stochastic process – called the state noise (resp. the measurement or observation noise) – modelling the random neuronal fluctuations that drive the resting state activity. In the observation equations, $\varphi$ are the unknown parameters of the (haemodynamic) observation function and $u(t)$ represents any exogenous (or experimental) inputs that drive the hidden states, which are usually absent in resting state designs (Friston et al., 2014).

Spectral DCM furnishes a constrained inversion of the stochastic model by parameterising the neuronal fluctuations $v(t)$. Spectral DCM simplifies the generative model by replacing the original timeseries with their second-order statistics (i.e., cross spectra). This means that, instead of estimating time varying hidden states, we are estimating their covariance, which is time invariant. Then we simply need to estimate the covariance of the random fluctuations, where a scale free (power law) form for the state noise (resp. observation noise) is used, motivated from previous work on neuronal activity (Beggs & Plenz, 2003; Shin & Kim, 2006; Stam & De Bruin, 2004), as follows:

$$g_v(\omega, \theta) = \alpha_v \omega^{-\beta_v}$$
$$g_e(\omega, \theta) = \alpha_e \omega^{-\beta_e} \quad \text{(S3)}$$

Here, $\{\alpha, \beta\} \subset \theta$ are the parameters controlling the amplitudes and exponents of the spectral density of the neural fluctuations. The parameterisation of endogenous fluctuations means that the states are no longer probabilistic; hence the inversion scheme is significantly simpler, requiring estimation of only the parameters (and hyperparameters) of the model.

We used standard Bayesian model inversion to infer the parameters of the model in (1), (2) and (3), from the observed signal $y(t)$. The description of the Bayesian model inversion procedures based on variational Laplace can be found elsewhere (Friston et al., 2007; Friston et al., 2003; Razi & Friston, 2016).

**Parametric Empirical Bayes**

Empirical Bayes refers to the Bayesian inversion or fitting of hierarchical models. In hierarchical models, constraints on the posterior density over model parameters at any given level are provided by the level above. These constraints are called empirical priors because they are informed by empirical data. A hierarchical Parametric Empirical Bayes (PEB) model for DCM parameters has recently been introduced, which represents how individual (within-subject) connections derive from the subjects' group membership (Friston et al., 2016). Mathematically, for DCM studies with *N* subjects and *M* parameters per DCM, we have a hierarchical model, where the responses of the *i*-th subject and the distribution of the parameters over subjects can be modeled as:

$$y_i = \Gamma_i^{(1)}(\theta^{(1)}) + \varepsilon_i^{(1)}, \qquad \text{(S4)}$$
$$\theta^{(1)} = \Gamma^{(2)}\left(\theta^{(2)}\right) + \varepsilon^{(2)} ,$$
$$\theta^{(2)} = \eta + \varepsilon^{(3)} ,$$

where, $y_i$ is the BOLD time series from *i-th* subject *and* $\Gamma_i^{(1)}$ is a nonlinear mapping from the parameters of a model to the predicted response $y$, which in this study was the model in Eq. S1 above. $\varepsilon_i^{(1)}$ is independent and identically distributed (i.i.d.) observation noise (equivalent to $e(t)$ in Eq. S2). In this hierarchical form, *empirical priors* encoding second (between-subject) level effects place constraints on subject-specific parameters. The second level is a linear model where the random effects are parametrised in terms of their precision:

$$\Gamma^{(2)}\left(\theta^{(2)}\right) = (X \otimes W)\beta , \qquad \text{(S5)}$$

where, $\beta \subset \theta$ are group means or effects encoded by a design matrix with between-subject, $X$, and within-subject, $W$, parts. The between-subject part encodes differences among subjects or covariates such as age, while the within-subject part specifies mixtures of parameters that show random effects. We assume that the first column of the design matrix is a constant term, modelling group means, and subsequent columns encode group differences.

Table S1. Summary of Group Differences in Fronto-striato-thalamic Effective Connectivity in FEP (n = 46), FEP-SCZ (n = 17), and SCZ (n = 36) Patients Respective to Healthy Controls

| Connection | Increased (+) or decreased (-) in patients | Effect size (Hz) | 90% Posterior Confidence Interval (lower bound, upper bound) | Patients (mean) | Controls (mean) |
|---|---|---|---|---|---|
| **FEP vs HCs** | | | | | |
| *dlPFC → dlPFC* | + | *0.10* | *0.03, 0.20* | *0.64* | *0.44* |
| Thal → NAcc | - | 0.10 | -0.20, -0.01 | -0.12 | 0.09 |
| Amyg → NAcc | + | 0.11 | 0.02, 0.21 | 0.00 | -0.24 |
| *VTA/SN → VTA/SN* | - | *0.14* | *-0.24, -0.04* | *-0.04* | *0.00* |
| | | | | *(-0.48)* | *(-0.05)* |
| **FEP-SZ vs HCs** | | | | | |
| Thal → NAcc | - | 0.13 | -0.24, -0.03 | -0.13 | 0.09 |
| Amyg → NAcc | + | 0.13 | 0.02, 0.23 | 0.00 | -0.25 |
| *Amyg → Amyg* | - | *0.11* | *-0.22, 0.00* | *-0.16* | *0.00* |
| | | | | *(-0.43)* | *(-0.05)* |
| NAcc → Hipp | - | 0.15 | -0.26, -0.04 | -0.28 | 0.00 |
| VTA/SN→ Hipp | - | 0.09 | -0.20, -0.02 | -0.14 | 0.00 |
| VTA/SN→ NAcc | + | 0.09 | -0.01, 0.20 | 0.00 | -0.14 |
| *VTA/SN→ VTA/SN* | - | *0.08* | *-0.19, 0.03* | *-0.06** | *0.07** |
| | | | | *(-0.47)* | *(-0.54)* |
| **SCZ vs HCs** | | | | | |
| dlPFC → Thal | - | 0.11 | -0.19, -0.03 | -0.14 | 0.00 |
| vmPFC → dlPFC | - | 0.08 | -0.16, 0.00 | -0.06* | 0.06* |
| Thal → Hipp | - | 0.10 | -0.18, -0.02 | -0.20 | 0.00 |
| Thal → NAcc | - | 0.09 | -0.17, -0.01 | -0.11 | 0.00 |
| Amyg → Hipp | - | 0.09 | -0.17, -0.01 | 0.00 | 0.16 |
| *VTA/SN → VTA/SN* | - | *0.16* | *-0.25, -0.07* | *-0.26* | *-0.09* |
| | | | | *(-0.39)* | *(-0.46)* |
| VTA/SN → DC | - | 0.14 | -0.22, -0.06 | -0.19 | 0.00 |
| DC → VTA/SN | + | 0.16 | 0.08, 0.25 | 0.12 | -0.11 |

* denotes mean connections that were derived from the DCM PEB routine as these connections were removed in the subsequent Bayesian model averaging routine.

All connections have the posterior probability value of 1.00

All parameters for between region connections are in Hz.

Self-connections are italicized, and values are log-transformed to ensure prior negativity (i.e., inhibitory) constraints on self-connections. A positive value for self-connection denotes increased inhibition, a negative value signifies reduced inhibition. Self-connection parameters in Hz are displayed in parentheses.

Table S2. Summary of Connections Associated with Severity of Clinical Symptoms in FEP (n = 46), FEP-SCZ (n = 17), and SCZ (n = 36) Patients

| Connection | Positive (+) or negative (-) association | Effect size (Hz) | 90% Posterior Confidence Interval (lower bound, upper bound) |
|---|---|---|---|
| **Positive symptoms** | | | |
| **FEP** | | | |
| dlPFC → vmPFC | - | 0.10 | -0.20, 0.01 |
| *vmPFC → vmPFC* | - | *0.16* | *-0.27, -0.06* |
| *Amyg → Amyg* | - | *0.08* | *-0.19, 0.02* |
| Hipp → VTA/SN | + | 0.10 | 0.00, 0.20 |
| NAcc → VTA/SN | + | 0.09 | -0.01, 0.20 |
| VTA/SN → NAcc | + | 0.09 | -0.02, 0.20 |
| VTA/SN → Hipp | - | 0.17 | -0.27, -0.06 |
| VTA/SN → Amyg | + | 0.14 | 0.04, 0.25 |
| VTA/SN → DC | - | 0.08 | -0.19, 0.02 |
| **FEP-SCZ** | | | |
| *vmPFC → vmPFC* | - | *0.19* | *-0.33, -0.05* |
| *dlPFC → dlPFC* | - | *0.09* | *-0.23, 0.04* |
| dlPFC → Thal | + | 0.11 | -0.02, 0.24 |
| *Thal → Thal* | + | *0.13* | *0.00, 0.26* |
| Hipp → Amyg | - | 0.15 | -0.28, -0.02 |
| NAcc → Hipp | - | 0.09 | -0.22, 0.03 |
| NAcc → VTA/SN | + | 0.12 | -0.01, 0.25 |
| DC → Thal | - | 0.12 | -0.25, 0.02 |
| VTA/SN → NAcc | + | 0.13 | 0.01, 0.26 |
| VTA/SN → Hipp | - | 0.14 | -0.27, -0.01 |
| VTA/SN → Amyg | + | 0.10 | -0.03, 0.23 |
| VTA/SN → DC | - | 0.12 | -0.25, 0.01 |
| **SCZ** | | | |
| *vmPFC → vmPFC* | - | *0.09* | *-0.20, 0.03* |
| vmPFC → Hipp | + | 0.16 | 0.05, 0.27 |
| vmPFC → VTA/SN | + | 0.08 | -0.03, 0.20 |
| *dlPFC → dlPFC* | + | *0.09* | *-0.03, 0.21* |
| dlPFC → Thal | - | 0.18 | -0.24, -0.06 |
| Thal → Hipp | + | 0.11 | 0.00, 0.22 |
| Thal → VTA/SN | - | 0.13 | -0.24, -0.02 |
| Hipp → vmPFC | - | 0.09 | -0.21, 0.02 |

| | | | |
|---|---|---|---|
| DC → Thal | + | 0.08 | -0.03, 0.19 |
| *VTA/SN → VTA/SN* | - | *0.15* | *-0.27, -0.04* |
| VTA/SN → Amyg | - | 0.10 | -0.21, 0.01 |
| VTA/SN → dlPFC | + | 0.11 | 0.00, 0.21 |
| **Negative symptoms** | | | |
| **FEP** | | | |
| *dlPFC → dlPFC* | + | *0.11* | *0.01, 0.21* |
| Amyg → Thal | - | 0.09 | -0.19, 0.02 |
| Amyg → Hipp | + | 0.08 | 0.02, 0.19 |
| *Hipp → Hipp* | + | *0.12* | *0.02, 0.22* |
| DC → VTA/SN | - | 0.09 | -0.20, 0.01 |
| VTA/SN → Thal | - | 0.13 | -0.24, -0.03 |
| **FEP-SCZ** | | | |
| vmPFC → Amyg | - | 0.19 | -0.32, -0.05 |
| *dlPFC → dlPFC* | + | *0.11* | *-0.02, 0.24* |
| dlPFC → Thal | + | 0.12 | -0.01, 0.26 |
| dlPFC → DC | + | 0.09 | -0.06, 0.24 |
| Thal → Amyg | + | 0.17 | 0.03, 0.30 |
| *Amyg → Amyg* | + | *0.14* | *0.00, 0.27* |
| Amyg → Thal | - | 0.11 | -0.23, 0.02 |
| *Hipp → Hipp* | + | *0.14* | *0.01, 0.27* |
| **SCZ** | | | |
| *Thal → Thal* | - | *0.17* | *-0.28, -0.06* |
| Thal → vmPFC | + | 0.09 | -0.02, 0.19 |
| *Amyg → Amyg* | - | *0.16* | *-0.27, -0.05* |
| *NAcc → NAcc* | - | *0.09* | *-0.20, 0.03* |
| NAcc → Hipp | - | 0.10 | -0.22. 0.01 |
| NAcc → VTA/SN | + | 0.11 | -0.01, 0.22 |
| *VTA/SN → VTA/SN* | + | *0.10* | *-0.01, 0.21* |
| VTA/SN → Hipp | - | 0.13 | -0.24, -0.02 |
| VTA/SN → dlPFC | - | 0.11 | -0.22, 0.00 |

All connections have the posterior probability (free energy) value of 1.00

All parameters for between region connections are in Hz. Self-connections are italicized, and values are log-transformed to ensure prior negativity (i.e., inhibitory) constraints on self-connections. A positive value for self-connection denotes increased inhibition, a negative value signifies reduced inhibition.

Positive and negative symptoms measured with BPRS Positive and Negative subscales.

Table S3. Summary of Connections Associated with Striatal Dopamine Synthesis

| Connection | Positive (+) or negative (-) association | Effect size (Hz) | 90% Posterior Confidence Interval (lower bound, upper bound) |
|---|---|---|---|
| **Dorsal Striatum** | | | |
| *Thal → Thal* | + | *0.10* | *-0.04, 0.23* |
| Thal → DC | + | 0.09 | -0.04 0.22 |
| Thal → VTA/SN | + | 0.12 | -0.01, 0.26 |
| Amyg → vmPFC | + | 0.08 | -0.05, 0.21 |
| Amyg → Thal | + | 0.11 | -0.02, 0.24 |
| DC → Thal | - | 0.13 | -0.26, 0.00 |
| **Ventral Striatum** | | | |
| Hipp → Thal | + | 0.16 | 0.03. 0.30 |
| Hipp → VTA/SN | + | 0.16 | 0.03, 0.29 |
| Amyg → Thal | - | 0.12 | -0.25, 0.02 |
| NAcc → Thal | - | 0.11 | -0.24, 0.02 |
| *Amyg → Amyg* | - | *0.14* | *-0.27, 0.01* |
| *NAcc → NAcc* | - | *0.14* | *-0.28, 0.00* |
| DC → VTA/SN | - | 0.12 | -0.25, 0.01 |
| *DC → DC* | + | *0.10* | *-0.03, 0.22* |
| *VTA/SN → VTA/SN* | + | *0.10* | *-0.05, 0.25* |
| VTA/SN → vmPFC | + | 0.15 | 0.02, 0.29 |
| VTA/SN → DC | + | 0.12 | -0.01, 0.25 |

All connections have the posterior probability value of 1.00

All parameters for between region connections are in Hz. Self-connections are italicized, and values are log-transformed to ensure prior negativity (i.e., inhibitory) constraints on self-connections. A positive value for self-connection denotes increased inhibition, a negative value signifies reduced inhibition.

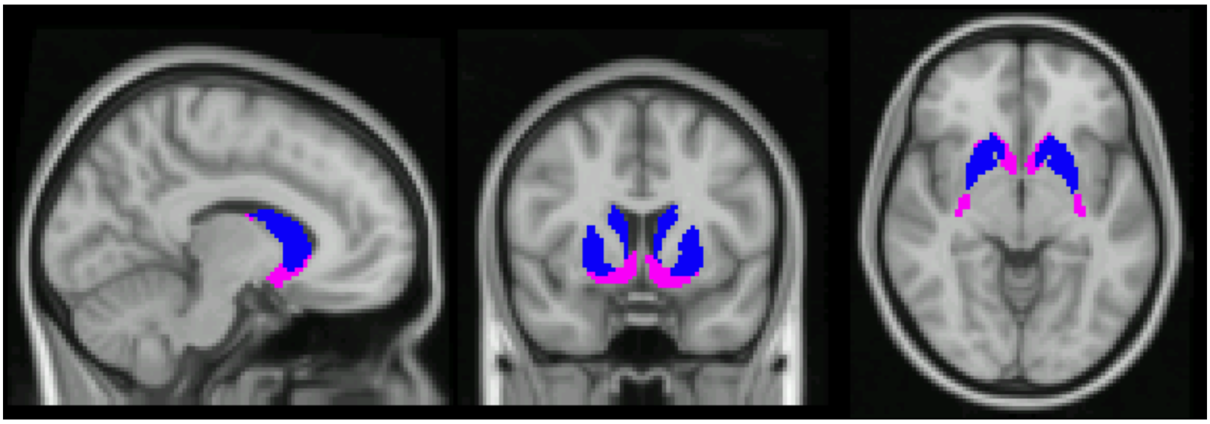

**Figure S1. Parcellation of Dorsal and Ventral Striatal Regions of Interest in PET Quantification Presented on the MNI 2mm Template.** The ventral striatum is in magenta and dorsal striatum is in blue. Striatal ROIs were registered to each person's anatomical template. PET analysis was restricted to the left hemisphere.

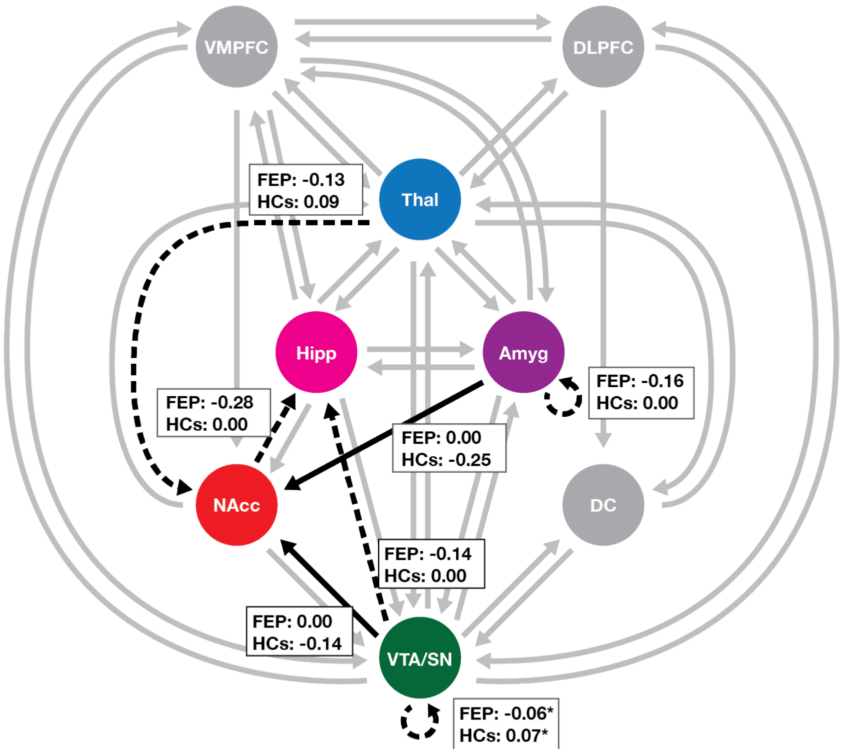


**Figure S2. Group Differences in Effective Connectivity Parameters between FEP-SCZ Patients (n = 17) and Healthy Controls (n = 23).** Boxes show mean connectivity values in each group. Colors of regions in the top panel correspond with the colors in connectivity diagrams. For connections between regions, dashed arrows represent connections for which patients show a greater inhibitory influence compared to controls; solid arrow represents connections for which patients show greater excitatory, or less inhibitory, influence compared to controls. For self-connections, dashed arrows represent reduced inhibition (i.e., lower negative values) in patients compared to controls. Gray arrows represent modeled connections that were not (significantly) different from the prior. All connectivity parameters are in Hz, including self-connections. Connections were thresholded at Pp > 0.95.

* denotes mean connections that were derived from the DCM PEB routine as these connections were removed in the subsequent Bayesian model averaging routine.

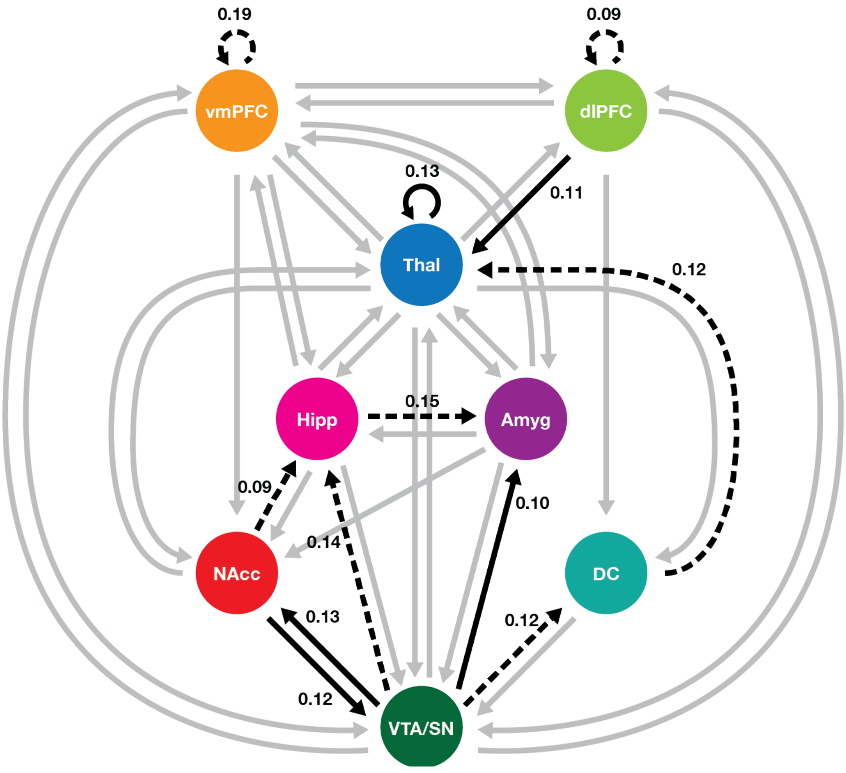


**Figure S3. Effective Connectivity Associated with Positive Symptoms in FEP-SCZ Patients (n = 17).** Solid arrows: positive associations between effective connectivity parameters and positive PLEs; dashed arrows: negative associations between effective connectivity parameters and positive PLEs; gray arrows: associations that were not (significantly) different from the prior. Connections were thresholded at Pp > 0.95.

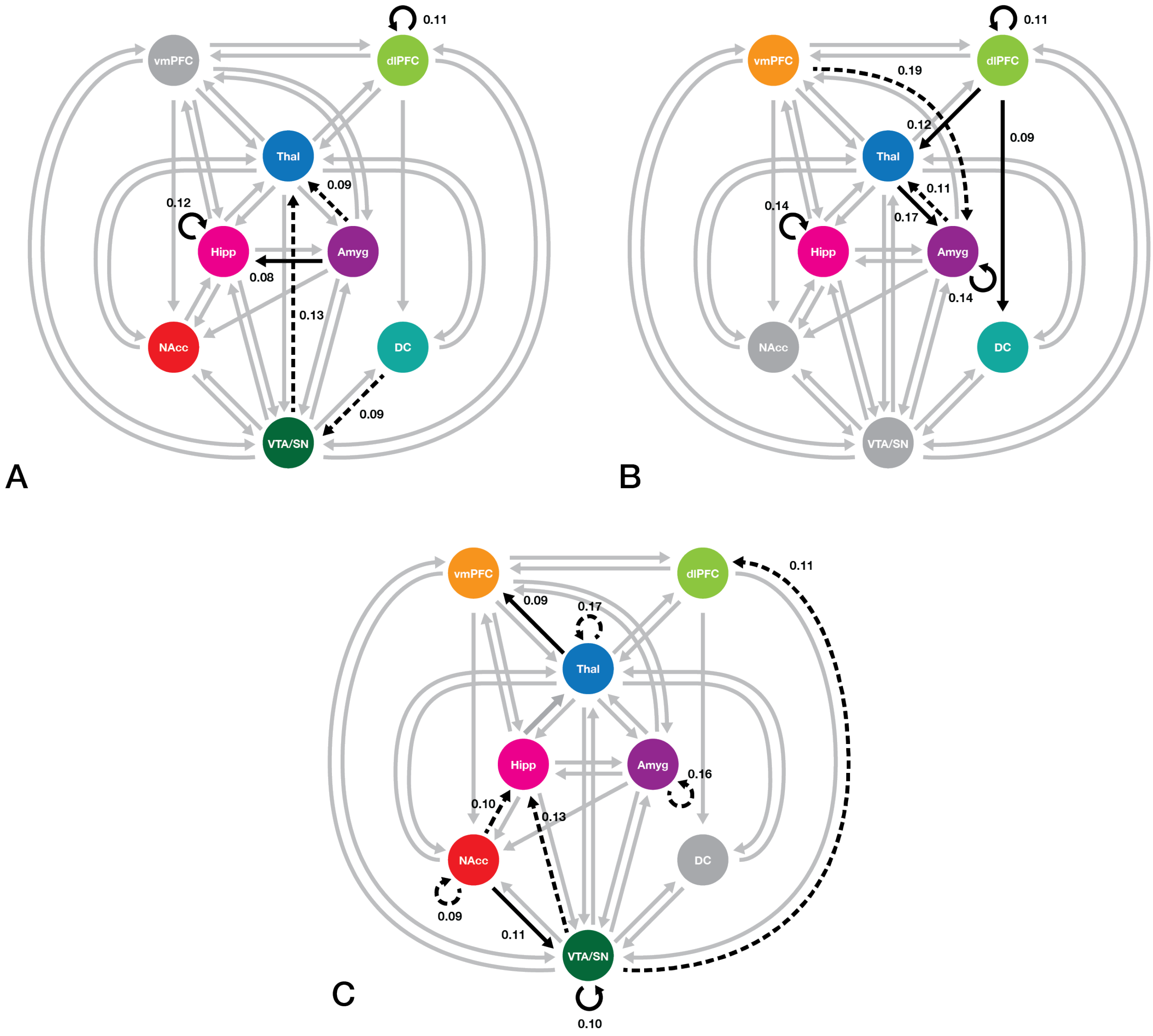


**Figure S4. Associations Between Severity of Negative Symptomatology and Effective Connectivity Parameters Across All Cohorts**. Panels depict associations in (A) FEP (n = 46), (B) FEP-SCZ (n = 17), and (C) SCZ (n = 17). Solid arrows: positive associations between effective connectivity parameters and negative PLEs/symptoms; sashed arrows: negative associations between effective connectivity parameters and negative PLEs/symptoms; gray arrows: associations that were not (significantly) different from the prior. Connections were thresholded at Pp > 0.95.
